# Chloroquine and hydroxychloroquine effectiveness in human subjects during coronavirus: a systematic review

**DOI:** 10.1101/2020.05.07.20094326

**Authors:** Salman Rawaf, Mohammed N Al-Saffar, Harumi Quezada-Yamamoto, Mashael Alshaikh, Michael Pelly, David Rawaf, Elizabeth Dubois, Azeem Majeed

**Author notes:** CORRESPONDING AUTHOR: Professor Salman Rawaf, Department of Primary Care and Public Health, School of Public Health, Faculty of Medicine, Imperial College London, Reynolds Building, St Dunstan’s Road, London, W6 8RP.

## Abstract

In a search to find effective treatments for COVID-19, chloroquine and hydroxychloroquine have gained attention. We aim to provide evidence to support clinical decision-making regarding medication for the treatment of COVID-19 by carrying out a systematic review of the literature. The electronic databases MEDLINE, EMBASE, Global Health, and HMIC were searched up to April 2020. Eligible study outcomes included: extubation or patient recovery. Relevant data were extracted and analysed by narrative synthesis. Our results included six studies in the review of which four studies were of good or fair quality. All eligible studies included were for coronavirus involving the use of either chloroquine or hydroxychloroquine to treat common symptoms such as fever, cough, shortness of breath and fatigue. Outcomes most commonly reported were improved lung function, viral clearance, and hospital discharge. Strong evidence to support the use of chloroquine and hydroxychloroquine in the treatment of COVID-19 is lacking. Fast track trials are riddled with bias and may not conform to rigorous guidelines which may lead to inadequate data being reported. The use of these drugs in combination with other medications may be useful but without knowing which groups they are suited for and when they may cause more harm than good.

## Introduction

As the COVID-19 pandemic has streaked around the plant, the pursuit for therapeutic options has developed at a fast pace. Coronaviruses are not new. In the past two decades, the virus was responsible for previous outbreaks of Severe Acute Respiratory Syndrome (SARS) and the Middle East Respiratory Syndrome (MERS). Yet despite this experience, no clear treatment pathway had been agreed in some countries. Therefore, this current pandemic of a variant novel virus has taken the world by surprise with the only option of delivering empirical treatment at the early stages, until a vaccine is available.

In a search to find effective treatments for COVID-19, chloroquine (CQ) and hydroxychloroquine have entered the spotlight (1). Current evidence comes from poorly controlled clinical trials demonstrating antiviral activity against severe-acute respiratory syndrome coronavirus 2 (SARS-CoV-2) (2). Systematic reviews of variable quality have started to appear focusing on current patients without looking at past evidence with other viruses of the same family (3). To date, no systematic reviews have been published examining the clinical effectiveness of chloroquine and hydroxychloroquine in the context of the current pandemic or of past treatment for patients with severe coronavirus respiratory infections.

Past outbreaks of coronaviruses have documented some useful treatments including chloroquine and hydroxychloroquine. These compounds are used to treat malaria, systemic lupus erythematosus and other rheumatic diseases. Chloroquine increases endosomal pH required for virus/cell fusion and interferes with the glycosylation of cellular receptors of SARS-CoV (4). Authors Wang *et al*. (5) reported that chloroquine functions at both entry and post-entry stages of the 2019-nCoV and in addition to its antiviral activity, has an immune-modulating effect (5). The 90% effective concentration (EC90) value of chloroquine against the 2019-nCoV in vitro, was demonstrated to be clinically achievable in the plasma of rheumatoid arthritis patients who received 500 mg (6). The metabolism of chloroquine after oral administration occurs mostly in the liver. Its excretion is slow and maintains a plasma half-life of 2.5 to 10 days. Furthermore, individuals with impaired or compromised liver function at baseline (e.g. ventilated patients in ITU with multiple fat-soluble infusions running) are more likely to experience accumulation in-vivo and require close monitoring of liver function test and risk of liver failure. The adult acute lethal dose of chloroquine is between two to four grams in ages 18 to 65, according to the Wuhan Institute of Virology (7).

The study does not to stop at what medication is appropriate but also requires knowing when it is better to start treatment. From SARS we know that clinical worsening of individuals in Week 2 is apparently more related to immunopathological damage than to uncontrolled coronavirus replication (8). Keyaerts *et al*. (9) observed that chloroquine displayed significant anti-SARS-CoV activity (9), but that inhibitory capability sharply declined if not administered within five-hour post infection (9). Yet, advantages of chloroquine such as low cost and well-established safety could allow its use as prophylaxis in individuals at high risk such as healthcare workers (10).

The aim of this research is to report the existent clinical evidence of chloroquine and hydroxychloroquine effectiveness, either alone or in combination, in the recovery of human patients infected with coronavirus respiratory infections. In addition, difference in dosages and treatment initiation times will be analysed.

## Materials and Methods

### Literature search

Literature searches with medical electronic databases were conducted for studies published from 1950 onwards: Ovid MEDLINE, EMBASE, Global Health, and HMIC. Please refer to S1 file for an example of our search strategy.

### Eligibility criteria

Studies on the use of chloroquine and hydroxychloroquine in treatment for coronavirus respiratory symptoms, on human patients (children or adults) diagnosed with SARS, MERS, COVID-19. Studies needed to include at least one of the following outcomes: elimination of active infection (detected in blood or swabs), recovery understood as no active infection or reduction of symptoms to an acceptable level for discharge or extubation from ventilators. Only studies with full text available in English were included. Studies conducted solely in healthy subjects or for the common cold were excluded, as were rapid reviews, narrative reviews, comments, opinion pieces, methodological reports, editorials, letters and conference abstracts. Non-human studies such as mice or in-vitro cultures were also excluded. The search included MeSH terms.

### Screening and selection

Study selection was conducted by two reviewers independently. Title and abstract screening followed by full texts were performed using *Covidence* software against eligibility criteria. After deduplication, each reviewer summarised results and compared. Any disagreement was resolved by discussion. Discrepancies were resolved by consensus. This systematic review was conducted according to the Preferred Reporting Items for Systematic Reviews and Meta-Analysis (PRISMA) guidelines (Please see S1 file: Table 4).

### Data extraction

Selected studies were exported, stored and tracked on the computer software reference manager Zotero. Data relevant to the study question were extracted from included studies and summarized. Information on author, study design, associated with the treatment of coronavirus using chloroquine or hydroxychloroquine was collected.

### Assessment of study quality and risk bias

The quality of the primary studies was assessed by three reviewers and scored using the National Heart, Lung, and Blood Institute (NHLBI) quality assessment tools for controlled intervention studies, observational studies, and systematic reviews (11). For quality assessment in case reports and case series, Murad *et al. (12)* tool was used. Studies were not excluded based on quality assessment. Studies were critically appraised for risk of bias.

### Outcome measures

Outcomes such as extubation from ventilators or patient recovery. The latter defined as no active infection in either blood or swabs; or reduction of symptoms to an acceptable level for patient discharge from hospital.

### Data synthesis and analysis

Due to methodological heterogeneity and varying clinical outcome measures reported across studies, a meta-analysis of results was not performed. A narrative synthesis of the finding was conducted.

## Results

### Characteristics of selected studies

The search identified 575 papers, of which six studies met the eligibility criteria (Please see S2 file: Figure 1): two systematic reviews (13,14), one randomised control trial (15), one non-randomised clinical trial (16), one an observational cohort study (17), and one case report (18). Study characteristics are summarised in Table 1.

**Figure 1.**
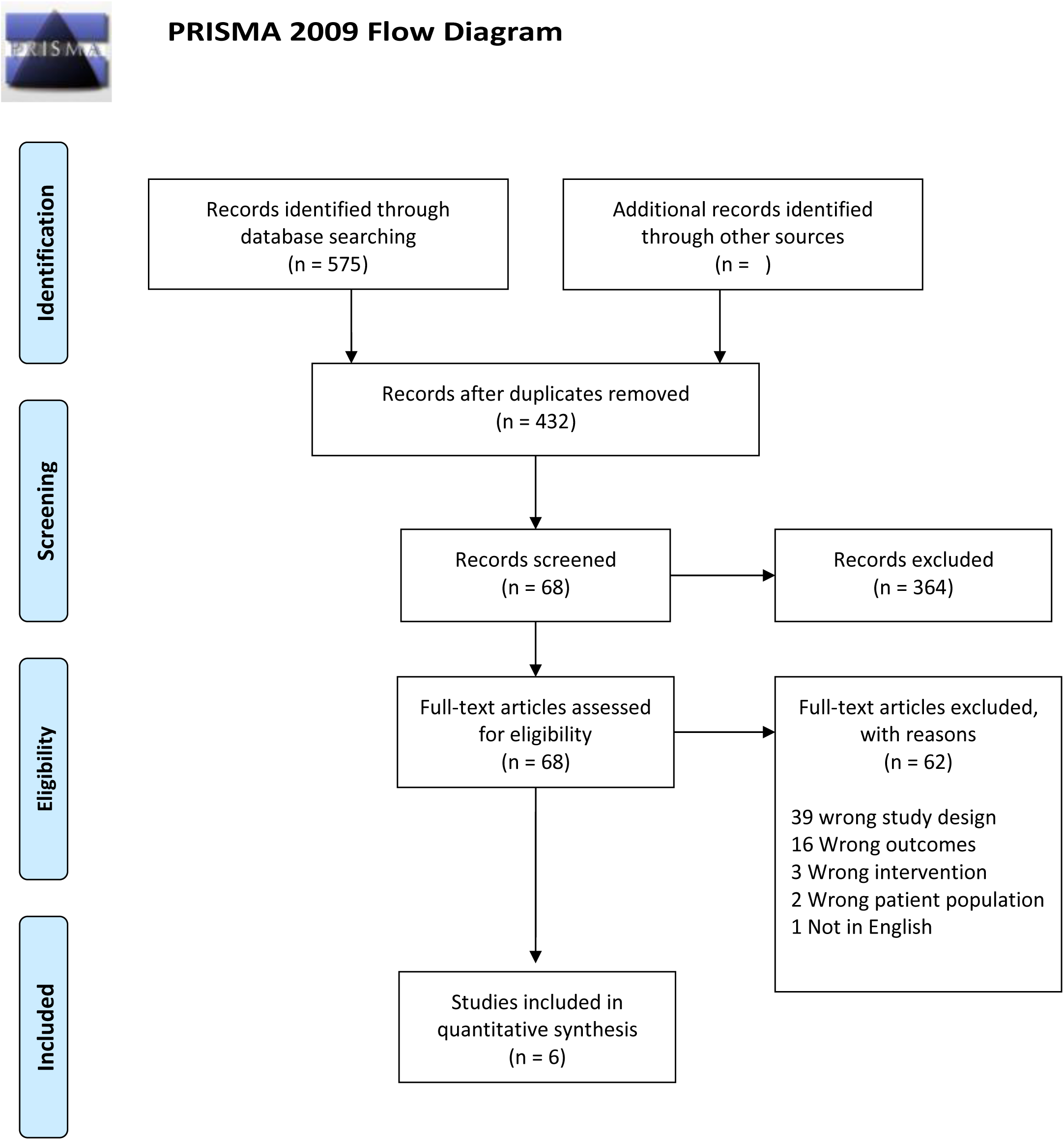
PRISMA flow diagram of search strategy showing different phases of the selection process.

**Table 1.** Characteristics of studies on the treatment of coronavirus using chloroquine and hydroxychloroquine.

| **Author** | **Type of study** | **Tool** | **Score** |
| --- | --- | --- | --- |
| Spezzani V *et al.* (15) | Case report | Murad *et al.* Tool for evaluating the methodological quality of case reports and case series | 5/6 (originally 8 items but 2 NA)<br>83%<br>Good |
| Sarma P *et al.* (11) | Systematic review and Meta-analysis | NHLBI Quality Assessment of Systematic Reviews and Meta-Analyses | 7/8<br>87%<br>Good. |
| Gautret P *et al.* (14) | Uncontrolled non-comparative observational study (cohort) | NHLBI Quality Assessment Tool for Observational Cohort and Cross-Sectional Studies | 7/13 (originally 14 items but 1 NA)<br>57%<br>Fair |
| Singh A K *et al.* (10) | Systematic Review | NHLBI Quality Assessment of Systematic Reviews and Meta-Analyses | 3/7 (originally 8 items but 1 NA)<br>42%<br>Poor |
| Huang M *et al.* (12) | Randomised control trial | NHLBI Quality Assessment of Controlled Intervention Studies | 7/14<br>50%<br>Fair |
| Gautret P *et al.* (13) | Open-label non-randomised controlled clinical trial | NHLBI Quality Assessment of Controlled Intervention Studies | 4/12 (originally 14 items but 2 NA)<br>33%<br>Poor |

### Quality and risk bias of selected studies

The six selected studies were scored using the National Heart, Lung, and Blood Institute (NHLBI, Maryland, USA) and Murad *et al* quality assessment tools. Two scored poor (13,16); two as good (14,18); and two as fair (15,17) (Table 1).

### Effectiveness of chloroquine and hydroxychloroquine in treatment of coronavirus

The clinical study by Huang *et al*. (15) demonstrated that patients treated with chloroquine (500 mg orally, twice daily for 10 days) appear to show better patient recovery compared with those patients treated with lopinavir/ritonavir. As a result, the patients treated with chloroquine were discharged from hospital earlier. Table 1 summarises the results of eligible studies on the effectiveness of drugs in treating infected coronavirus patients.

In the study by Gautret *et al*. (16) 70% of patients treated with 600 mg of hydroxychloroquine (200 mg, three times per day for 10 days) were virologically cured at day six post inclusion, compared to 12.5% of patients in the control group (p=0.001). In another group, 100% of Patients treated with hydroxychloroquine and azithromycin were virologically cured at day 6 post inclusion compared with 57.1% patients treated with hydroxychloroquine alone, and 12.5% in the control group (p<0.001). Gautret *et al*. (17) carried out a cohort study where they looked at the outcomes of patients treated with a combination of hydroxychloroquine sulfate (200 mg, three times per day for 10 days for four days) and antibiotic azithromycin (500 mg on day 1 followed by 250 mg per day for next four days), reporting positive results from the study. A broad-spectrum antibiotic (ceftriaxone) was added in those who developed pneumonia.

The case report study by Spezzani *et al*. (18) reported that patients treated with darunavir/cobicistat and hydroxychloroquine (200 mg, twice daily) in combination with a triple antibiotic therapy (levofloxacin, piperacillin plus tazobactam) had a better outcome compared to darunavir/cobicistat and hydroxychloroquine combined with double therapy of ceftriaxone and azithromycin. Both Italian patients started treatment at admission, seven days after initial symptoms. Despite this, the couple achieved remission on different weeks as the course of the disease differed due to individual risk factors. Patient one had metastatic breast cancer and recent exposure to antineoplastic chemotherapy which had produced leukopenia (immunosuppression) at admission, whereas there was no hint of a significant immunosuppression of patient two. However, patient one had a rapid recovery compared to a prolonged and more severe course compared to patient two who had a relatively low risk profile except for hypertension.

The systematic review by Singh *et al*. (13) looked at the effects of hydroxychloroquine and its impact on COVID-19 patients with type 2 diabetes in resource constrained settings with reference to India. They provide the current dosage guidelines on chloroquine and hydroxychloroquine use from China, South Korea, United States, Netherlands, Canada, and Belgium for the treatment of COVID-19 using chloroquine and hydroxychloroquine (13). Dosage recommendations for adults from each of these sources vary depending on the severity of the cases. Based on the results of the study, the authors conclude that because of its limited side effects, availability, and cost-effectiveness, the drugs should be worthy for fast track clinical trials for treatment of COVID-19. However, another systematic review by Sarma *et al*. (14) found that when compared to conventional treatment, there was no difference observed in virological cure, death, clinical worsening of disease, or safety. The main benefit was that treatment with hydroxychloroquine alone resulted in a lower number of cases showing radiological progression of lung disease. Additional benefits included less days to temperature normalisation and lowered total cough days compared to conventional treatment. The authors recommended that more data is acquired before making a definitive conclusion on the safety and effectiveness of the drugs.

## Discussion

The results of this systematic review indicate a positive trend favouring the use of chloroquine singularly or the combination of hydroxychloroquine with antibiotic therapy (regardless of added bacterial infection at the beginning of the treatment). Evidence was insufficient to favour a treatment start on Week one versus Week two (or vice versa) of symptoms appearing. However, Spezzani *et al*. (18) showed that immunosuppression may actually enhance treatment effectiveness by the use of the combination of hydroxychloroquine, antibiotic therapy and darunavir/cobicistat in patients who started treatment seven days after initial symptoms.

These findings have implications for clinical practice and policy in the current pandemic. Despite the potential therapeutic effect of chloroquine and hydroxychloroquine, fears exist that excess demand may lead to a shortage for people with other diseases who are currently taking these drugs (19).

Chloroquine and hydroxychloroquine are usually safe and well tolerated in normal dosage but can be extremely toxic in overdose. Potential adverse effects that should be considered before prescribing include prolongation of the QT interval (especially in pre-existing cardiac disease or if combined with azithromycin), hypo-glycemia, neuropsychiatric effects, drug–drug interactions and idiosyncratic hypersensitivity reactions (20). Moreover, chloroquine is not as widely available as hydroxychloroquine in some countries and is associated with greater adverse effects such as interaction with lopinavir/ritonavir, resulting in prolongation of the QT interval (21).

### Strengths and limitations

To our knowledge, this systematic review is the first attempt to gather evidence from fully published studies that focus on the treatment, to date, of coronavirus outbreaks in human subjects. Contrasting to ours, previous research explores the suitability of either chloroquine or hydroxychloroquine in treating coronavirus by relying on findings from animal studies and dosage recommendations from unpublished trials. Our search identified six eligible studies. Two scored highly in the methodological quality assessment. This may be due to small sample size, unclear or absent randomisation, concealment, blinding, ambiguous research question and objectives to help readers understand the purpose of the studies.

### Comparison with existing literature

Two studies (15,16) outline key information on socio-demographic and clinical characteristics; both used comparison groups to test the effectiveness of the drugs. Patients were tested before hospital admission and then prior to being administered the specific dosage of chloroquine and hydroxychloroquine. In both studies, patients were monitored and given treatment for 10 days with reported outcomes focused on viral clearance and lung improvement. Our review also included a case-report (18) identifying two patients from the same household discharged from hospital following combination therapy of antibiotics and hydroxychloroquine (18).

The results found no previous research on treatments using hydroxychloroquine or chloroquine targeting coronavirus such as SARS and MERS, except for COVID-19. More recent studies try to highlight the mechanisms of COVID-19 in animal studies and in cell cultures. The most cited successful human subject trials regarding the effectiveness of hydroxychloroquine and chloroquine were from China and France, by Gao *et al*. (22) and Gautret *et al*. (16). A recent published review focused on understanding the effectiveness of hydroxychloroquine and chloroquine in treating COVID-19 (23), but includes articles that have not had their results formally published. These articles focus mostly on COVID-19 treatments (23) and do not consider work done previously on coronaviruses such as SARS and MERS. The publication by Gao *et al*. (22) merely provides a list of ongoing trials, which is why it did not meet the eligibility criteria for this review. Our review included two additional systematic reviews and a case-report which met our inclusion criteria (13,14,18). A summary of past and ongoing trials found across the included studies can be consulted in Table 2.

**Table 2.** Past and ongoing clinical trials for hydroxychloroquine and chloroquine in coronavirus

| Mentioned in | Cites | Country | Intervention | Outcome | Status | Highlights |
| --- | --- | --- | --- | --- | --- | --- |
| Singh A K et al | Gao et al | China | Case-control study<br>They only they tried it on 100 vs control<br>They used In this study, chloroquine was given in dose of 500 mg of chloroquine twice daily in mild to severe COVID-19 pneumonia | Chloroquine phosphate seems superior to the control treatment in inhibiting the exacerbation of pneumonia, improving lung imaging findings, promoting a virus negative conversion, and shortening the disease course according to the news briefing. | Ongoing | This trial is not yet published and only available in a letter form, interestingly, this early result led China to include chloroquine in the prevention and treatment of COVID-19 pneumonia |
| Singh A K et al | Gautret et al (included in this systematic review) | France, Marseille, | Open-label, non-randomized trial (n = 36) oral hydroxychloroquine sulfate 200 mg, three times per day during ten days. A total of 26 patients received hydroxychloroquine and 16 were control patients | Hcq alone and combination of Hcq plus azithromycin was highly and significantly effective in clearing viral nasopharyngeal carriage (measured by polymerase chain reaction [PCR]) in only three-to six days in COVID-19 subjects, compared to control | First results | The virological clearance day-6 post-inclusion (primary outcome) with Hcq vs. control was 70.0% versus 12.5%, respectively (p = 0.001).<br>Note: a small sample size, dropout of six patients and limited follow-up, apart from the non-randomized and open-label nature of the trial. |
| Gautret P et al. | Gautret et al (included in this systematic review) | France, Marseille, | Uncontrolled, non-comparative, observational study (n=80)<br>Combination of 200mg of oral hydroxychloroquine sulfate, three times per day for ten days combined with azithromycin (500mg on day 1 followed by 250 | The majority (65/80, 81.3%) of patients had favourable outcome and were discharged from the unit. Only 15% required oxygen therapy during their stay in our ID ward. | First results | Three patients were transferred to the ICU, of whom two improved and were then returned to the ID ward. One 74 year-old patient was still in ICU at the time of writing. Finally, one 86 year-old patient who was not transferred to the ICU, died in the ID ward |
|  |  |  | mg per day for the next four days).<br>For patients with pneumonia and NEWS score≥5, a broad-spectrum antibiotic (ceftriaxone) was added to hydroxychloroquine and azithromycin. |  |  |  |
| Sarma P et al. | Chen et al | China, Shanghai | 30 treatment naive patients with confirmed COVID-19<br>Hydroxychloroquine (n=15) Patients in HCQ group were given HCQ 400 mg per day for 5 days plus conventional symptomatic treatments<br>Control (n=15). Conventional symptomatic treatment alone. | No difference in viral cure between the two groups on day 7. | Not available | Article full-text available only in Chinese. |
| Sarma P et al. | Zhaowei et al, 2020 | China, Wuhan | 62 patients with confirmed COVID -19 Patients in the HCQ treatment group received additional oral HCQ (hydroxychloroquine sulfate tablets) 400 mg/d (200 mg/bid) between days 1 and 5<br>Control: Standard treatment (oxygen therapy, antiviral agents, antibacterial agents, and immunoglobulin, with or without corticosteroids). | HCQ treatment decreased time to body temperature normalization and number of cough days compared to control.<br>Less number of patients in the HCQ arm showed evidence of radiological progression. | First results | This article is a preprint and has not been peer-reviewed. It reports new medical research that has yet to be evaluated and so should *not* be used to guide clinical practice. |
| Sarma P et al. | Million et al | France,<br>Marseille | 1061 COVID -19 patients<br>treated for minimum 3<br>days with HCQ+<br>Azithromycin combination<br>No control | Virological cure on 10th Day:<br>91.7%. Mortality: 0.47%. Total<br>cured till publication of study<br>report=98 % | First<br>results | Poor clinical outcomes were described for 4.3% of the patients, including<br>five death (0.5%). |

HCQ= hydroxychloroquine

### Implications for research and practice

At this stage of the pandemic, it is difficult to debate the treatment options for COVID-19. There are many ideas and theories being considered about this novel virus and currently clinicians lack the necessary evidence to effectively treat the infection. There may be a genetic influence with regards to susceptibility to the virus. However, contracting COVID-19 is multifactorial and it is important to investigate the exposure and host factors which put people at risk of infection. For example, it is unknown at what dosage the infectious particles begin to overwhelm the host’s immune system. This may inadvertently affect those in close contact with asymptomatic individuals. Thus, inadvertent exposure is a strong risk factor for infection. In such cases, social distancing may mitigate infection. Age, sex, ethnicity, and socio-economic backgrounds also need to be considered as potential risk factors when sampling from the population. Understanding the impact comorbidities such as cardiovascular disease, diabetes, hypertension, and obesity have on coronavirus infection is important in finding suitable treatments for COVID-19.

Recent public and media attention in many countries on the use chloroquine and hydroxychloroquine has increased focus on repurposing the drugs to combat the COVID-19 pandemic. This has prompted the World Health Organisation to reconsider leaving both drugs out from a large trial to study the effectiveness and safety of promising medications suitable for treating COVID-19 patients (24). Other institutions have also began launching fast track trials to understand whether they help in the recovery time and outcomes, but these types of studies come with issues of design bias which is unlikely to provide important data on the true effects of the drugs. Without essential data to provide key information about the suitability of these compounds in different populations, it will difficult to provide them to those who need them the most. Future research should adhere to the rigorous standard guidelines for the randomised control and observational cohort studies as best as possible, so that valuable and unbiased information is provided on these medications.

## Conclusion

The current evidence that exists on real human patients is weak despite effectiveness shown in in-vitro cultures for past coronavirus outbreaks and with the COVID-19 variant. It is unclear if there is an effect on the effectiveness, depending on early or late stage of administration. Nevertheless, recent clinical trials suggest a more positive outcome for those patients treated with chloroquine singularly or hydroxychloroquine combinations. Off-label use of these drugs for COVID-19 could raise the demand which would require a counterbalance in production. Otherwise, this may lead to a negative impact for those treated for malaria, lupus and other rheumatic diseases. Further randomised trials are needed urgently.

## Data Availability

All data are fully available without restriction

## Acknowledgements

This research was supported by the National Institute for Applied Research Collaboration (ARC) for Northwest London (NIHR ARC NWL). The views expressed in this article are those of the author(s) and not necessarily those of the NHS, the NIHR, or the Department of Health and Social Care.

## Author contributions

Salman Rawaf (SR) conceived the study. Mohammed Al-Saffar (MAS) and Harumi Quezada-Yamamoto (HQY) initiated study design, protocol, analysed the data, drafted the original manuscript and interpreted the results. Elizabeth Dubois (ED), HQY and MAS selected studies for inclusion and assessed quality. Mashael Alshaikh (MA), ED, HQY and MAS, extracted data, and revised subsequent drafts. Michael Pelly (MP), David Rawaf (DR), Azeem Majeed (AM), HQY and ED added specific critical review, commentary or revision. All authors discussed data analyses and contributed to interpretation of results. All authors approved the final manuscript for publication. The guarantor is Salman Rawaf.

## Conflict of interest

None declared

---

**Supplemental Table 1.** Study characteristics of studies meeting eligibility criteria for data synthesis.

| **Author** | **Type of study** | **Tool** | **Score** |
| --- | --- | --- | --- |
| Spezzani V *et al.* (15) | Case report | Murad *et al.* Tool for evaluating the methodological quality of case reports and case series | 5/6 (originally 8 items but 2 NA)<br>83%<br>Good |
| Sarma P *et al.* (11) | Systematic review and Meta-analysis | NHLBI Quality Assessment of Systematic Reviews and Meta-Analyses | 7/8<br>87%<br>Good. |
| Gautret P *et al.* (14) | Uncontrolled non-comparative observational study (cohort) | NHLBI Quality Assessment Tool for Observational Cohort and Cross-Sectional Studies | 7/13 (originally 14 items but 1 NA)<br>57%<br>Fair |
| Singh A K *et al.* (10) | Systematic Review | NHLBI Quality Assessment of Systematic Reviews and Meta-Analyses | 3/7 (originally 8 items but 1 NA)<br>42%<br>Poor |
| Huang M *et al.* (12) | Randomised control trial | NHLBI Quality Assessment of Controlled Intervention Studies | 7/14<br>50%<br>Fair |
| Gautret P *et al.* (13) | Open-label non-randomised controlled clinical trial | NHLBI Quality Assessment of Controlled Intervention Studies | 4/12 (originally 14 items but 2 NA)<br>33%<br>Poor |

**Supplemental Table 2.** Past and ongoing clinical trials for hydroxychloroquine and chloroquine in coronavirus

| Mentioned in | Cites | Country | Intervention | Outcome | Status | Highlights |
| --- | --- | --- | --- | --- | --- | --- |
| Singh A K et al | Gao et al | China | Case-control study<br>They only they tried it on 100 vs control<br>They used In this study, chloroquine was given in dose of 500 mg of chloroquine twice daily in mild to severe COVID-19 pneumonia | Chloroquine phosphate seems superior to the control treatment in inhibiting the exacerbation of pneumonia, improving lung imaging findings, promoting a virus negative conversion, and shortening the disease course according to the news briefing. | Ongoing | This trial is not yet published and only available in a letter form, interestingly, this early result led China to include chloroquine in the prevention and treatment of COVID-19 pneumonia |
| Singh A K et al | Gautret et al (included in this systematic review) | France, Marseille, | Open-label, non-randomized trial (n = 36) oral hydroxychloroquine sulfate 200 mg, three times per day during ten days. A total of 26 patients received hydroxychloroquine and 16 were control patients | HCQ alone and combination of HCQ plus azithromycin was highly and significantly effective in clearing viral nasopharyngeal carriage (measured by polymerase chain reaction [PCR]) in only three-to six days in COVID-19 subjects, compared to control | First results | The virological clearance day-6 post-inclusion (primary outcome) with HCQ vs. control was 70.0% versus 12.5%, respectively (p = 0.001).<br>Note: a small sample size, dropout of six patients and limited follow-up, apart from the non-randomized and open-label nature of the trial. |
| Gautret P et al. | Gautret et al (included in this systematic review) | France, Marseille, | Uncontrolled, non-comparative, observational study (n=80)<br>Combination of 200mg of oral hydroxychloroquine sulfate, three times per day for ten days combined with azithromycin (500mg on day 1 followed by 250 mg per day for the next four days).<br>For patients with pneumonia and NEWS score ≥ 5, a broad- spectrum antibiotic (ceftriaxone) was added to hydroxychloroquine and azithromycin. | The majority (65/80, 81.3%) of patients had favourable outcome and were discharged from the unit. Only 15% required oxygen therapy during their stay in our ID ward. | First results | Three patients were transferred to the ICU, of whom two improved and were then returned to the ID ward. One 74 year-old patient was still in ICU at the time of writing. Finally, one 86 year-old patient who was not transferred to the ICU, died in the ID ward |
| Sarma P et al. | Chen et al | China, Shanghai | 30 treatment naive patients with confirmed COVID-19<br>Hydroxychloroquine (n=15) Patients in HCQ group were given HCQ 400 mg per day for 5 days plus conventional symptomatic treatments<br>Control (n=15). Conventional symptomatic treatment alone. | No difference in viral cure between the two groups on day 7. | Not available | Article full-text available only in Chinese. |
| Sarma P et al. | Zhaowei et al, 2020 | China, Wuhan | 62 patients with confirmed COVID -19 Patients in the HCQ treatment group received additional oral HCQ (hydroxychloroquine sulfate tablets) 400 mg/d (200 mg/bid) between days 1 and 5<br>Control: Standard treatment (oxygen therapy, antiviral agents, antibacterial agents, and immunoglobulin, with or without corticosteroids). | HCQ treatment decreased time to body temperature normalization and number of cough days compared to control.<br>Less number of patients in the HCQ arm showed evidence of radiological progression. | First results | This article is a preprint and has not been peer-reviewed. It reports new medical research that has yet to be evaluated and so should *not* be used to guide clinical practice. |
| Sarma P et al. | Million et al | France, Marseille | 1061 COVID -19 patients treated for minimum 3 days with HCQ+ Azithromycin combination<br>No control | Virological cure on 10th Day: 91.7%. Mortality: 0.47%. Total cured till publication of study report=98 % | First results | Poor clinical outcomes were described for 4.3% of the patients, including five death (0.5%). |

HCQ= hydroxychloroquine

## Appendix 1.

**Search terms:**

Studies included were last searched on the 19th April 2020.

The articles identified through the search included text words, in the following combination:

1. hydroxychloroquine.mp. or exp Hydroxychloroquine/
2. chloroquine.mp. or exp Chloroquine/
3. exp Coronavirus/ or Coronavirus.mp. or exp Coronavirus Infections/
4. MERS.mp. or exp Middle East Respiratory Syndrome Coronavirus/
5. Middle East Respiratory Syndrome.mp.
6. exp SARS Virus/ or SARS.mp. or exp Severe Acute Respiratory Syndrome/
7. Severe Acute Respiratory Syndrome.mp.
8. exp Respiratory Distress Syndrome, Adult/ or ARDS.mp.
9. Acute Respiratory Distress Syndrome.mp.
10. COVID.mp.
11. coronavirus 19.mp.
12. 1 or 2
13. 3 or 4 or 5 or 6 or 7 or 8 or 9 or 10 or 11
14. 12 and 13
15. remove duplicates from 14

## Appendix 2.

Preferred Reporting Items for Systematic Review and Meta-Analysis (PRISMA) Checklist

| Section/topic | # | Checklist item | Reported on page # |
| --- | --- | --- | --- |
| **TITLE** |  |  |  |
| Title | 1 | Identify the report as a systematic review, meta-analysis, or both. | 1 |
| **ABSTRACT** |  |  |  |
| Structured summary | 2 | Provide a structured summary including, as applicable: background; objectives; data sources; study eligibility criteria, participants, and interventions; study appraisal and synthesis methods; results; limitations; conclusions and implications of key findings; systematic review registration number. | 2 |
| **INTRODUCTION** |  |  |  |
| Rationale | 3 | Describe the rationale for the review in the context of what is already known. | 3, 4 |
| Objectives | 4 | Provide an explicit statement of questions being addressed with reference to participants, interventions, comparisons, outcomes, and study design (PICOS). | 3, 4 |
| **METHODS** |  |  |  |
| Protocol and registration | 5 | Indicate if a review protocol exists, if and where it can be accessed (e.g., Web address), and, if available, provide registration information including registration number. | 4 |
| Eligibility criteria | 6 | Specify study characteristics (e.g., PICOS, length of follow-up) and report characteristics (e.g., years considered, language, publication status) used as criteria for eligibility, giving rationale. | 5 |
| Information sources | 7 | Describe all information sources (e.g., databases with dates of coverage, contact with study authors to identify additional studies) in the search and date last searched. | 5 |
| Search | 8 | Present full electronic search strategy for at least one database, including any limits used, such that it could be repeated. | 5 |
| Study selection | 9 | State the process for selecting studies (i.e., screening, eligibility, included in systematic review, and, if applicable, included in the meta-analysis). | 6 |
| Data collection process | 10 | Describe method of data extraction from reports (e.g., piloted forms, independently, in duplicate) and any processes for obtaining and confirming data from investigators. | 6 |
| Data items | 11 | List and define all variables for which data were sought (e.g., PICOS, funding sources) and any assumptions and simplifications made. | 6 |
| Risk of bias in individual studies | 12 | Describe methods used for assessing risk of bias of individual studies (including specification of whether this was done at the study or outcome level), and how this information is to be used in any data synthesis. | 6 |
| Summary measures | 13 | State the principal summary measures (e.g., risk ratio, difference in means). | 6 |
| Synthesis of results | 14 | Describe the methods of handling data and combining results of studies, if done, including measures of consistency (e.g., I<sup>2</sup>) for each meta-analysis. | 6 |
| Section/topic | # | Checklist item | Reported on page # |
| Risk of bias across studies | 15 | Specify any assessment of risk of bias that may affect the cumulative evidence (e.g., publication bias, selective reporting within studies). | NA |
| Additional analyses | 16 | Describe methods of additional analyses (e.g., sensitivity or subgroup analyses, meta-regression), if done, indicating which were pre-specified. | NA |
| **RESULTS** |  |  |  |
| Study selection | 17 | Give numbers of studies screened, assessed for eligibility, and included in the review, with reasons for exclusions at each stage, ideally with a flow diagram. | 7 |
| Study characteristics | 18 | For each study, present characteristics for which data were extracted (e.g., study size, PICOS, follow-up period) and provide the citations. | 7 |
| Risk of bias within studies | 19 | Present data on risk of bias of each study and, if available, any outcome level assessment (see item 12). | 7 |
| Results of individual studies | 20 | For all outcomes considered (benefits or harms), present, for each study: (a) simple summary data for each intervention group (b) effect estimates and confidence intervals, ideally with a forest plot. | 7, 8, 9 |
| Synthesis of results | 21 | Present results of each meta-analysis done, including confidence intervals and measures of consistency. | NA |
| Risk of bias across studies | 22 | Present results of any assessment of risk of bias across studies (see Item 15). | NA |
| Additional analysis | 23 | Give results of additional analyses, if done (e.g., sensitivity or subgroup analyses, meta-regression [see Item 16]). | NA |
| **DISCUSSION** |  |  |  |
| Summary of evidence | 24 | Summarize the main findings including the strength of evidence for each main outcome; consider their relevance to key groups (e.g., healthcare providers, users, and policy makers). | 10 |
| Limitations | 25 | Discuss limitations at study and outcome level (e.g., risk of bias), and at review-level (e.g., incomplete retrieval of identified research, reporting bias). | 10, 11 |
| Conclusions | 26 | Provide a general interpretation of the results in the context of other evidence, and implications for future research. | 13 |
| **FUNDING** |  |  |  |
| Funding | 27 | Describe sources of funding for the systematic review and other support (e.g., supply of data); role of funders for the systematic review. | 14 |

## Notes

### Competing Interest Statement

The authors have declared no competing interest.

### Funding Statement

No external funding was received

